# Final sizes and durations of new COVID-19 pandemic waves in Ukraine and around the world predicted by generalized SIR model

**DOI:** 10.1101/2021.11.22.21266683

**Authors:** Igor Nesteruk

## Abstract

New waves of the COVID-19 pandemic in Ukraine, which began in the summer of 2021, and after holidays in the middle of October 2021, were characterized by almost exponential growth of smoothed daily numbers of new cases. This is a matter of great concern and the need to immediately predict the epidemic dynamics in order to assess the possible maximum values of new cases, the risk of infection and the number of deaths. The generalized SIR-model and corresponding parameter identification procedure was used to simulate and predict the dynamics of two new epidemic waves in Ukraine and one in the whole world. Results of calculations show that new cases in Ukraine will not stop to appear before November 2022. If the global situation with vaccination, testing and treatment will not change, the pandemic could continue for another ten years.

## Introduction

The COVID-19 pandemic dynamics in Ukraine was discussed in [1-14]. To predict the first wave of the pandemic, the classical SIR model [15-17] and the statistics-based method of its parameter identification [18] were used. To simulate new epidemic waves, a numerical method of their detection [3, 19], a generalized SIR-model [20], and a corresponding parameter identification procedure [21] were developed. In particular, eleven epidemic waves were simulated for Ukraine [4, 7-10] and five pandemic waves for the whole world [4].

The calculations of the 11th pandemic wave (based on the accumulated numbers cases reported by Ukrainian national statistics [22, 23] in the period May 23 – June 5, 2021) predicted the end of this wave on August 25, 2021 with the number of cases 2,226,797 (see [10]). As of August 25, 2021 the real number of cases accumulated in Ukraine was 2,278,171 (see Table 1). It means that the predicted saturation level was exceeded only 2.26% (after 81 days of observation). The obtained high accuracy of the method allows us to hope for a fairly accurate forecast for next pandemic waves in Ukraine (12th and 13th) and in the whole world (6th), to which this study is devoted. Some results concerning the 12th epidemic wave in Ukraine are already available in [13].

**Table 1.** Cumulative numbers of laboratory-confirmed Covid-19 cases and deaths in Ukraine in the summer and autumn of 2021 according to the national statistics, [22, 23].

| Day in corresponding month of 2021 | Number of cases in July, $V_j$ | Number of cases in August, $V_j$ | Number of cases in September, $V_j$ | Number of cases in October, $V_j$ | Number of cases in November, $V_j$ | Number of deaths in October, $d_j$ | Number of deaths in November $d_j$ |
| --- | --- | --- | --- | --- | --- | --- | --- |
| 1 | 2236497 | 2253534 | 2290848 | 2447222 | 2955693 | 56649 | 68727 |
| 2 | 2237202 | 2254361 | 2293541 | 2455189 | 2979086 | 56775 | 69447 |
| 3 | 2237579 | 2255345 | 2296155 | 2460010 | 3006463 | 56889 | 70146 |
| 4 | 2237823 | 2256397 | 2297534 | 2469856 | 3032951 | 57206 | 70842 |
| 5 | 2238364 | 2257478 | 2298307 | 2482518 | 3058014 | 57526 | 71635 |
| 6 | 2238974 | 2258532 | 2300504 | 2497643 | 3075433 | 57840 | 72084 |
| 7 | 2239591 | 2259151 | 2303276 | 2514005 | 3088501 | 58081 | 72557 |
| 8 | 2240246 | 2259451 | 2306939 | 2529913 | 3107489 | 58331 | 73390 |
| 9 | 2240753 | 2260232 | 2310554 | 2541257 | 3130772 | 58463 | 74206 |
| 10 | 2241043 | 2261354 | 2314423 | 2550089 | 3155519 | 58700 | 74857 |
| 11 | 2241217 | 2262601 | 2316619 | 2562085 | 3179577 | 59052 | 75601 |
| 12 | 2241698 | 2263864 | 2317824 | 2578394 | 3203149 | 59523 | 76302 |
| 13 | 2242245 | 2265217 | 2321156 | 2597275 | 3217639 | 59935 | 76705 |
| 14 | 2242868 | 2265912 | 2325796 | 2610899 | 3228441 | 60137 | 77147 |
| 15 | 2243605 | 2266329 | 2331540 | 2623882 | 3244749 | 60414 | 78085 |
| 16 | 2244196 | 2267219 | 2338164 | 2635170 | 3263417 | 60633 | 78754 |
| 17 | 2244495 | 2268666 | 2344398 | 2644694 | 3284008 | 60810 | 79506 |
| 18 | 2244677 | 2270226 | 2348381 | 2660273 | - | 61348 | - |
| 19 | 2245275 | 2271826 | 2350646 | 2679185 | - | 61843 | - |
| 20 | 2245930 | 2273558 | 2355805 | 2701600 | - | 62389 | - |
| 21 | 2246656 | 2274561 | 2362559 | 2725385 | - | 63003 | - |
| 22 | 2247419 | 2275171 | 2370425 | 2748614 | - | 63486 | - |
| 23 | 2248164 | 2275863 | 2379483 | 2769405 | - | 63872 | - |
| 24 | 2248450 | 2276590 | 2387750 | 2784039 | - | 64202 | - |
| 25 | 2248663 | 2278171 | 2392397 | 2803159 | - | 64936 | - |
| 26 | 2249344 | 2280203 | 2395404 | 2825733 | - | 65628 | - |
| 27 | 2250061 | 2282285 | 2401956 | 2851804 | - | 66204 | - |
| 28 | 2250907 | 2284191 | 2411622 | 2878674 | - | 66852 | - |
| 29 | 2251869 | 2284940 | 2423379 | 2904872 | - | 67393 | - |
| 30 | 2252785 | 2286296 | 2435413 | 2922302 | - | 67729 | - |
| 31 | 2253269 | 2288371 | - | 2936238 | - | 68027 | - |

### Data

We will use the data set regarding the accumulated numbers of laboratory-confirmed COVID-19 cases and deaths in Ukraine from national sources [22, 23]. The corresponding numbers *V*_*j*_, *d*_*j*_ and moments of time *t*_*j*_ (measured in days) are shown in Table 1 for the period of July to November 2021. The values *V*_*j*_, corresponding to the previous moments of time, can be found in [4, 8-10]. The period *T*_*c11*_: May 23 – June 5, 2021 has been used in [10] for SIR simulations of the eleventh epidemic wave in Ukraine. Here we use the datasets, corresponding to the period *T*_*c12*_: September 29 – October 12, 2021 to simulate the 12th wave and the period *T*_*c13*_: October 28 – November 10, 2021 for the 13th wave. Other *V*_*j*_ and *t*_*j*_ values will be used to control the accuracy of predictions.

To estimate the mortality rate in Ukraine (ratio of accumulated number of deaths *d*_*j*_ to accumulated number of cases *V*_*j*_), let us take figures *d*_*j*_ corresponding different days: 52,286 (June 26, 2021); 52,665 (July 13, 2021); 52,981 (August 2, 2021); 53,789 (August 30, 2021); 54,550 (September 15, 2021); 57,526 (October 5, 2021); 59,052 (October 11, 2021), [22, 23]. Taking corresponding *V*_*j*_ values we can calculate the mortality rates *m*_*i*_ =*d*_*j*_ *1000/*V*_*j*_ (per thousand of cases) for the listed days: 23.40; 23.49; 23.50; 23.52; 23.40; 23.17; 23.05. Thus, the mortality rate is rather stable (its variation during June to October 2021 is only 0.47). We will use the average value *m*= 23.36 to predict the number of deaths in Ukraine during the new 12th and 13th pandemic waves.

We will use the data set regarding the accumulated numbers of laboratory-confirmed COVID-19 cases in the whole world from the COVID-19 Data Repository by the Center for Systems Science and Engineering (CSSE) at Johns Hopkins University (JHU), [24]. The numbers *V*_*j*_ and moments of time *t*_*j*_ (measured in days) corresponding to the version of JHU data available on November 18, 2021 are shown in Table 2 for the period of May to November 2021. The period *T*_*c6*_: September 29 – October 12, 2021 will be used for SIR simulations of the sixth pandemic wave in the whole world. Other *V*_*j*_ and *t*_*j*_ values will be used to control the accuracy of predictions.

**Table 2.** Cumulative numbers of laboratory-confirmed Covid-19 cases in the whole world in the summer and autumn of 2021 according to the JHU datasets, [24].

| Day in corresponding month of 2021 | Number of cases in May, $V_j$ | Number of cases in June, $V_j$ | Number of cases in July, $V_j$ | Number of cases in August, $V_j$ | Number of cases in September $V_j$ | Number of cases in October, $V_j$ | Number of cases in November $V_j$ |
| --- | --- | --- | --- | --- | --- | --- | --- |
| 1 | 152276590 | 171272898 | 182726434 | 198444595 | 218595466 | 234356127 | 247171157 |
| 2 | 152951570 | 171758996 | 183167695 | 199022653 | 219272430 | 234701054 | 247599693 |
| 3 | 153630578 | 172248309 | 183544382 | 199660021 | 219990395 | 235008467 | 248117984 |
| 4 | 154438256 | 172667912 | 183872893 | 200335410 | 220474967 | 235452671 | 248643609 |
| 5 | 155280720 | 173067049 | 184242453 | 201024324 | 220914961 | 235872200 | 249157004 |
| 6 | 156150815 | 173390272 | 184695883 | 201846052 | 221355380 | 236384642 | 249569269 |
| 7 | 156983904 | 173710775 | 185159327 | 202397022 | 222080699 | 236816997 | 249914970 |
| 8 | 157770675 | 174077854 | 185639668 | 202838586 | 222712220 | 237290992 | 250391584 |
| 9 | 158412936 | 174497701 | 186147179 | 203492405 | 223349675 | 237622759 | 250871092 |
| 10 | 159031606 | 174947362 | 186574219 | 204141032 | 223983497 | 237937441 | 251442851 |
| 11 | 159770619 | 175368288 | 186944737 | 204869528 | 224439880 | 238319075 | 251956201 |
| 12 | 160530984 | 175741060 | 187381421 | 205580294 | 224808018 | 238751674 | 252542169 |
| 13 | 161256546 | 176044679 | 187901397 | 206385979 | 225409164 | 239214456 | 252974728 |
| 14 | 161974329 | 176352002 | 188442853 | 206919889 | 225960250 | 239658510 | 253318827 |
| 15 | 162603295 | 176723278 | 189014972 | 207383880 | 226527940 | 240115116 | 253860137 |
| 16 | 163152581 | 177121944 | 189613250 | 208062469 | 227104401 | 240449869 | 254382438 |
| 17 | 163691215 | 177514452 | 190087192 | 208745487 | 227697551 | 240764659 | - |
| 18 | 164313944 | 177921297 | 190518240 | 209473681 | 228234315 | 241178794 | - |
| 19 | 164986139 | 178270695 | 191014762 | 210189272 | 228594983 | 241619751 | - |
| 20 | 165265741 | 178573040 | 191544693 | 210979353 | 229128857 | 242090131 | - |
| 21 | 165892200 | 178866758 | 192103503 | 211519485 | 229597662 | 242546214 | - |
| 22 | 166471396 | 179238527 | 192667887 | 211965954 | 230135505 | 243030920 | - |
| 23 | 166948481 | 179676298 | 193357870 | 212670361 | 230646900 | 243390258 | - |
| 24 | 167400782 | 180081192 | 193829848 | 213357235 | 231194181 | 243705625 | - |
| 25 | 167933373 | 180503102 | 194275173 | 214087524 | 231560977 | 244135761 | - |
| 26 | 168502846 | 180869106 | 194815533 | 214823164 | 231909080 | 244576866 | - |
| 27 | 169057033 | 181180563 | 195425745 | 215568802 | 232381444 | 245088017 | - |
| 28 | 169557476 | 181508885 | 196070028 | 216116120 | 232830308 | 245545805 | - |
| 29 | 170039483 | 181888509 | 196722877 | 216562642 | 233332705 | 246049111 | - |
| 30 | 170431363 | 182285486 | 197453162 | 217249144 | 233817106 | 246429566 | - |
| 31 | 170810073 | - | 197963310 | 217868234 | - | 246749382 | - |

### Generalized SIR model and data smoothing procedure

The generalized SIR-model relates the number of susceptible *S*, infectious *I* and removed persons *R* for a particular epidemic wave *i*, [8, 20]. The exact solution of the set of non-linear differential equations uses the function

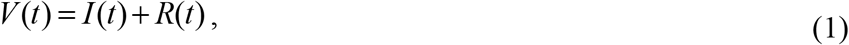

corresponding to the number of victims or the cumulative laboratory-confirmed number of cases [8, 20]. Its derivative:

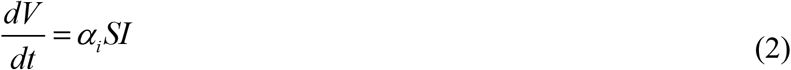

yields the estimation of the average daily number of new cases. When the registered number of victims *V*_*j*_ is a random realization of its theoretical dependence (1), the exact solution presented in [8, 20] depends on five parameters (α_*i*_ is one of them). The details of the optimization procedure for their identification can be found in [21].

Since daily numbers of new cases are random and characterized by some weekly periodicity, we will use the smoothed daily number of accumulated cases:

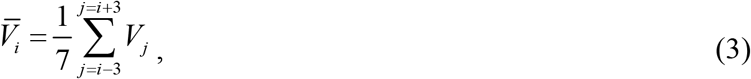

and its numerical derivative:

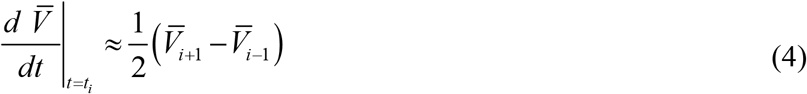

to estimate the smoothed number of new daily cases [3, 4, 9, 19].

## Results and discussion

The optimal values of the general SIR model and other characteristics of the 12th and 13th pandemic waves in Ukraine and the 6th wave in the whole world are calculated and listed in Table 3. The corresponding SIR curves are shown in Figs. 1 and 2 by blue and brown lines for Ukraine and green lines for the world. Black lines illustrate the results of SIR simulation of the eleventh epidemic wave in Ukraine published in [10]. It can be seen that the optimal values of SIR parameters are very different (even for 12th and 13th epidemic waves in Ukraine). Close values were obtained only for the average times of spreading the infection 1/ρ_*i*_. The assessments of the pandemic wave durations (corresponding the moment when the number of infectious persons becomes less that unit) are very pessimistic (November, 2022 for Ukraine and December 2031 for the whole world). A similar long epidemic wave was also predicted for India [25].

**Table 3.** Optimal values of parameters and other characteristics of the 12-th and 13-th COVID-19 pandemic waves in Ukraine and the 6-th wave in the whole world.

| Characteristics | 12<sup>th</sup> epidemic wave<br>in Ukraine,<br>$i=12$ | 13<sup>th</sup> epidemic wave<br>in Ukraine,<br>$i=13$ | 6<sup>th</sup> pandemic wave in<br>the whole world,<br>$i=6$ |
| --- | --- | --- | --- |
| Time period taken<br>for calculations $T_{ci}$ | September 29 –<br>October 12, 2021 | October 28 –<br>November 10, 2021 | September 29 –<br>October 12, 2021 |
| $I_i$ | 25,261.4089164122 | 73,492.9652436053 | 1,550,494.67573132 |
| $R_i$ | 2,399,050.73394073 | 2,801,190.17761354 | 231,782,051.609983 |
| $N_i$ | 3,790,400 | 7,237,600 | 334,411,200 |
| $\nu_i$ | 1,137,541.61656928 | 4,252,588.61675970 | 101736415.543763 |
| $\alpha_i$ | 2.63670321714285e-07 | 6.7868529452959e-08 | 2.84345358576434e-09 |
| $\rho_i$ | 0.299935964004210 | 0.288616935787874 | 0.289282775580724 |
| $1/\rho_i$ | 3.33404499630449 | 3.46480014164856 | 3.45682523956893 |
| $r_i$ | 0.997536473683354 | 0.998339758268927 | 0.999305866880929 |
| $S_{i\infty}$ | 845,264 | 3,503,575 | 84,981,994 |
| $V_{i\infty}$ | 2,945,136 | 3,734,025 | 249,429,206 |
| Final day of the<br>epidemic wave | June 16, 2022 | Nov 13, 2022 | December 2031 |

**Fig. 1.**
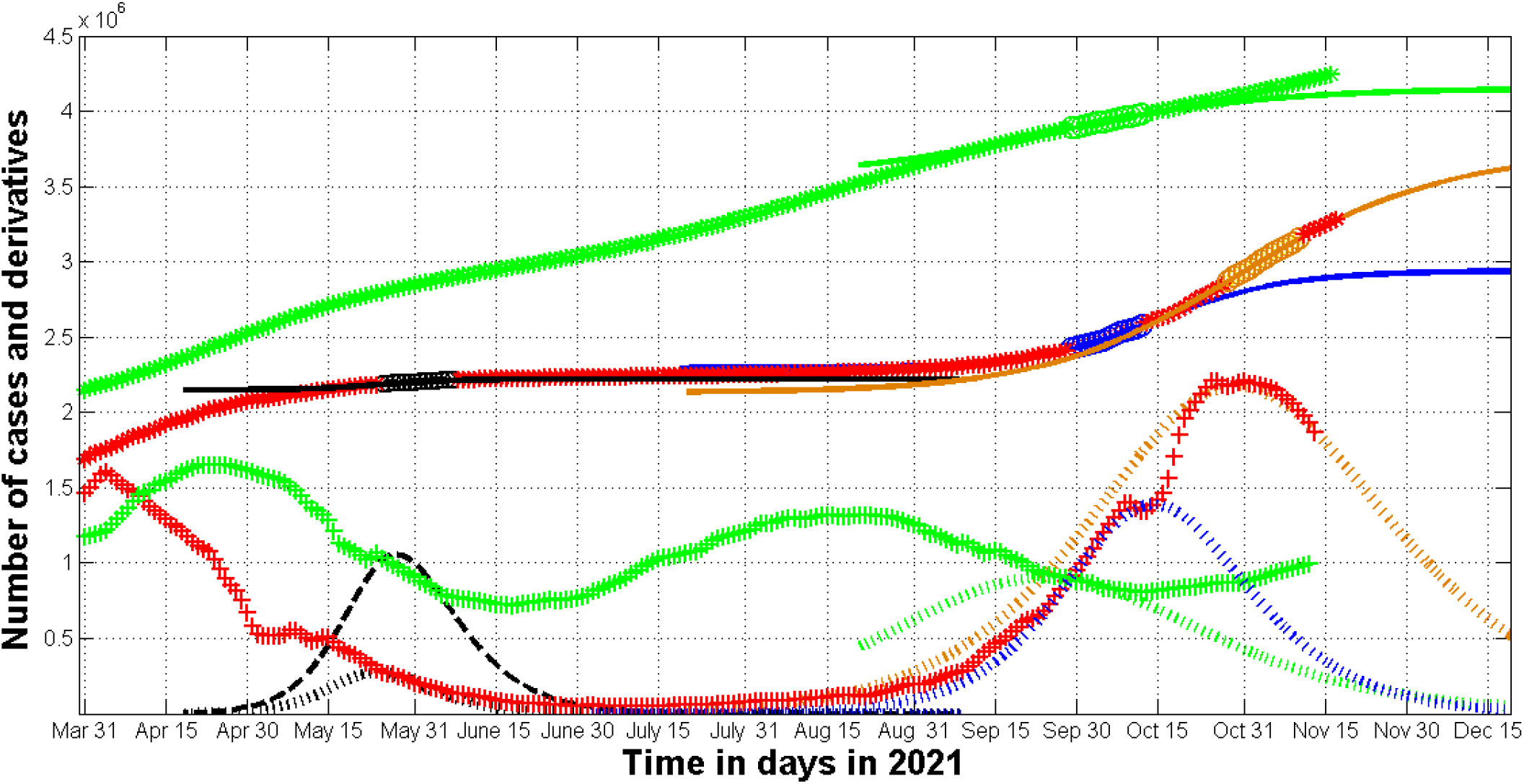
The COVID-19 pandemic waves in Ukraine and in the whole world in the summer and autumn of 2021. The results of SIR simulations of the 11th (see [10]), 12th, and 13th waves in Ukraine are shown by black, blue, and brown lines, respectively. Green lines represent the 6th pandemic wave in the whole world. Numbers of victims *V(t)=I(t)+R(t)* – solid lines (for the world divided by 60); numbers of infected and spreading *I(t)* multiplied by 5 – dashed; derivatives *dV/dt* (eq. (2), multiplied by 100 for Ukraine and by 2 for the world) – dotted. “Circles” correspond to the accumulated numbers of cases registered during the periods of time taken for SIR simulations (for the world divided by 60). “Stars” corresponds to *V*_*j*_ values beyond these time periods (for the world divided by 60). “Crosses” show the first derivative (4) multiplied by 100 for Ukraine and by 2 for the world.

**Fig. 2.**
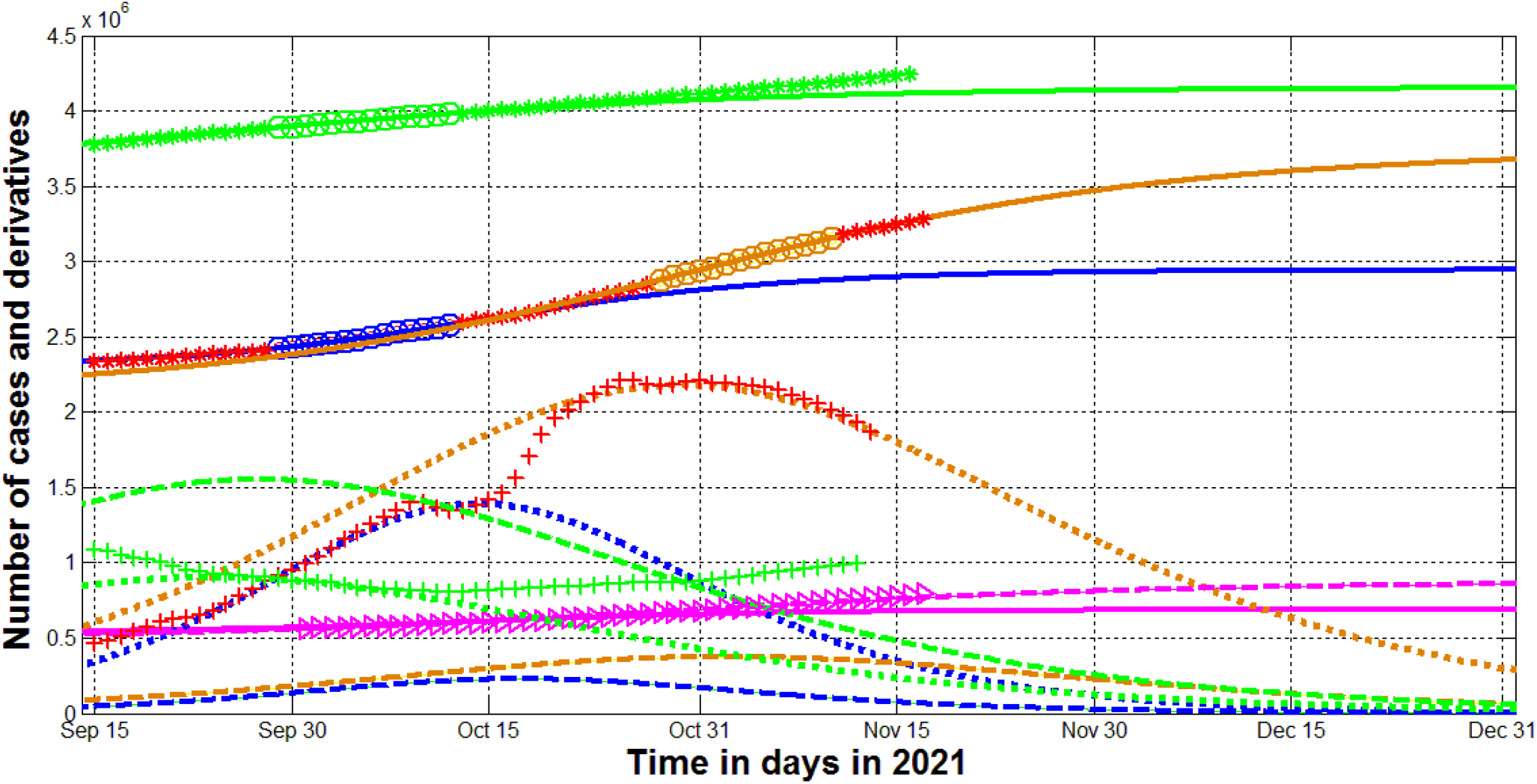
The COVID-19 pandemic waves in Ukraine and in the whole world in the autumn of 2021. The results of SIR simulations of the 12th and 13th waves in Ukraine are shown by blue and brown lines, respectively. Green lines represent the 6th pandemic wave in the whole world. Numbers of victims *V(t)=I(t)+R(t)* – solid lines (for the world divided by 60); numbers of infected and spreading *I(t)* (multiplied by 5 for Ukraine) – dashed; derivatives *dV/dt* (eq. (2), multiplied by 100 for Ukraine and by 2 for the world) – dotted. The magenta lines represent the estimation of the accumulated number of deaths during the 12th (solid) and 13th (dashed) epidemic waves in Ukraine multiplied by 10. Magenta “triangles” represent the accumulated numbers of death in Ukraine form Table 1 multiplied by 10. “Circles” correspond to the accumulated numbers of cases registered during the periods of time taken for SIR simulations (for the world divided by 60). “Stars” corresponds to *V*_*j*_ values beyond these time periods (for the world divided by 60). “Crosses” show the first derivative (4) multiplied by 100 for Ukraine and by 2 for the world.

The saturation levels (final sizes) *V*_*i∞*_ of the 12th wave in Ukraine and 6th global wave are already exceeded (compare corresponding values in Tables 1-3). As of November 17, 2021 the real accumulated number of deaths – 79,506 - registered in Ukraine (see Table 1) has already exceeded the figure 68,764 predicted in [13] for the end of 2021 with the use of *V(t)* curve for 12th epidemic wave. This discrepancy can be explained by the sharp increase in the daily number of new cases which occurred after long holidays October 14-17, 2021 (see red “crosses” in Figs. 1 and 2). These changes in the epidemic dynamics indicate the beginning of a new (13th) wave in Ukraine. The calculations allow us to estimate the new saturation level *V*_13∞_ = 3,734,025 (see Table 3) and the expected accumulated number of deaths 3,734,025^*^0.02336=87,227 by November 2022. Registered numbers of deaths in Ukraine agree with the theoretical estimation for 13th wave (compare the magenta “triangles” and the dashed magenta line in Fig. 2).

According to the predictions for the 12th wave (posted in [13]], the numbers of infectious persons and average daily new cases will stop to increase around 17 and 14 October 2021, respectively (see blue dashed and dotted lines in Fig. 2). The registered smoothed daily number of new cases in Ukraine really achieved a local maximum on October 10, 2021, but started to increase very rapid after October 17, 2022 (see the red “crosses” in Figs. 1 and 2).

“Stars” and “crosses” in Figs. 1 and 2 illustrate the accuracy of simulations for the accumulated number of cases and the averaged daily numbers of new cases (eq. (4)). Comparisons with corresponding blue solid and dotted lines in Fig. 2 show that the theoretical estimations for 12th wave in Ukraine were consistent with observations before October 15, 2021. After October 18 the results of observations are very close to the theoretical estimations for the 13-th wave (see brown solid and dotted lines in Fig. 2). As of November 17, 2021 the number of infectious persons (the brown dashed line) and the daily number of new cases were decreasing. Premature lifting of quarantine restrictions, a significant increase in contacts during the New Year and Christmas holidays or/and the appearance of a new coronavirus strain could disrupt these positive trends.

Unfortunately, the general SIR model cannot predict the emergence of new epidemic waves. It simulates the dynamics for only the period with constant epidemic conditions. Therefore, permanent monitoring of the number of new cases is needed to determine changes in the epidemic dynamics. After that it is possible to do new simulations by means of the generalized SIR model with calculation and use of new values of its parameters.

We can only point out the three possible reasons for the new 13th wave in Ukraine:

1. The long weekend of October 14-17, 2021 without significant quarantine restrictions led to a significant increase in travels and contacts. This period accounted for the maximum number of infected (see the blue dashed curve in Fig. 2). We observed a similar situation in Ukraine in May 2020, when the lockdown was lifted during the period of the maximum number of infectious people, which led to the emergence of the second epidemic wave before the end of the first one, [4, 9]. An increase in contacts during the holidays in early May 2021 also led to an increase in the number of infectious persons (see the black dashed line in Fig. 1). But during this period there was a tendency to reduce the daily number of new cases, so the increase in contacts only slowed down this trend (see red “crosses” in Fig. 1).
2. Due to a large number of asymptomatic patients, many COVID-19 cases are not detected and registered [26-31]. The ratio of real to detected cases in Ukraine was estimated to be between 4 and 20 for different periods of time [8, 10]. Such large numbers of undetected cases may suddenly change the number of reported cases, if the population frightened by the increase in mortality begins to seek medical care more often.
3. Appearance of new coronavirus strains. The global number of new cases is also characterized by wave-like behavior (see green “crosses” in Fig. 1). But unlike Ukraine and many other countries, the difference between the minimum and maximum values of the derivative (4) is much smaller for the world dynamics. The minima of new global cases also do not go to zero (compare green and red “crosses” in Figs. 1 and 2). All this limits the use of the SIR model for the long-term predictions. In particular, the increase in daily number of new cases (see green “crosses” in Figs. 1 and 2) indicate the beginning of a new global wave after October 15, 2021 (this fact makes the predictions for the 6^th^ wave no more relevant). It should be noted that the COVID-19 pandemic is characterized by a very slow decline in the number of infectious *I(t)*. In particular, according to the results of modeling of the 6th world wave (shown in Table 3), the number of infectious persons worldwide may be less than 100 in May 2021. This small number is enough to continue the pandemic for almost 10 years.

## Conclusions

The generalized SIR-model and corresponding parameter identification procedure was used to simulate and predict the dynamics of two new epidemic waves in Ukraine and one in the whole world. Results of calculations show that new cases in Ukraine will not stop to appear before November 2022. If the global situation with vaccination, testing and treatment will not change, the pandemic could continue for another ten years.

## Data Availability

All data produced in this study are available after reasonable request to the author

## Acknowledgements

The author is grateful to Oleksii Rodionov for his help in collecting and processing data.

